# Telehealth versus Self-Directed Lifestyle Intervention to Promote Healthy Blood Pressure: a Randomized Controlled Trial

**DOI:** 10.1101/2022.03.31.22273248

**Authors:** Alex R Chang, Lauren Gummo, Christina Yule, Heather Bonaparte, Charlotte Collins, Allison Naylor, Lawrence Appel, Stephen P Juraschek, Lisa Bailey-Davis

## Abstract

**Introduction:** Lifestyle behavior modification interventions, delivered using telehealth, have been shown to be effective in reducing weight. However, limited data exists on the benefits of lifestyle behavior change delivered using telehealth and web-based applications on blood pressure (BP).

**Methods:** We conducted a 2-site randomized controlled trial in the Geisinger Health System (January 2019 to March 2021) to compare the efficacy of a self-guided vs. a dietitian telehealth approach using web-based applications in 187 participants with 24-hour systolic BP 120-160 mmHg and body mass index ≥ 25 kg/m^2^. Both arms received recommendations to improve diet based on a web-based food frequency questionnaire, and access to an online weight management program. The telehealth arm received weekly telephone calls with a dietitian who used motivational interviewing. The primary outcome was 12-week change in 24-hour systolic BP. Secondary outcomes included changes in sleep/awake systolic BP and diastolic BP, self-reported physical activity, healthy eating index (HEI)-2015 score, and weight.

**Results:** A total of 187 participants (mean age 54.6 [SD 13.2] years, 52% female, 23% on BP medications, mean body mass index 34.5 [6.5] kg/m^2^, mean HEI-2015 score 60.8 [11.1] units) were randomized with 156 (83.4%) completing the trial. Mean 24-hour systolic BP improved from baseline to 12 weeks similarly in the dietitian (−6.73 mmHg, 95% CI: −8.64, −4.82) and the self-directed arm (−4.92, 95% CI: −7.01, −2.77; p comparing groups=0.2). The dietitian telehealth arm had greater 12-week improvements in sleep systolic BP (mean −6.92 vs. −1.45; p=0.004), sleep diastolic BP (−3.31 vs. 0.73; p=0.001), and self-reported physical activity (866 vs. −243 metabolic equivalent of task minutes/week; p=0.01). The dietitian telehealth arm also tended to have greater 12-week improvements in weight loss (−5.11 vs. −3.89 kg; p=0.1) and HEI-2015 score (9.23 vs. 6.43 units; p=0.09), though these differences were not statistically significant.

**Conclusions:** Dietitian-led telehealth supported by web-based applications resulted in a similar reduction in 24-hour systolic BP as a self-directed approach, with secondary improvements in sleep BP and physical activity.

**Trial registration number:** ClinicalTrials.gov Identifier NCT03700710

## Introduction

Hypertension is a leading cause of morbidity and mortality, affecting 121.5 million adults ≥20 years of age in the United States(1). Unhealthy dietary patterns, high sodium intake, low physical activity, and obesity contribute significantly to the pathogenesis of hypertension and cardiovascular diseases (2, 3). The American College of Cardiology/American Heart Association (ACC/AHA) 2017 Hypertension Guidelines recommend that patients with elevated systolic blood pressure (120-129 mmHg) and hypertension (≥130/80 mmHg) undergo lifestyle modification. Evidence from prior studies have demonstrated that weight loss, healthy dietary patterns (e.g. the Dietary Approaches to Stop Hypertension (DASH) or Mediterranean), and increased physical activity lower blood pressure (4–8).

Helping patients lower blood pressure through lifestyle modification is challenging in clinical practice as providers often lack time or resources to address this important need (9–11). Telehealth interventions with mobile health applications and online programs could be useful in aiding patients improve lifestyle behaviors and nutrition-related outcomes by providing insights based on their own data(12). Prior telehealth lifestyle behavior modification trials have been shown to be as effective as traditional, in-person interventions for weight reduction (13, 14). A recent pragmatic cluster-randomized clinical trial conducted at 15 Brigham and Women’s primary care practices examined the effectiveness of a combined intervention (online weight management program plus population health management) with use of the online program only, and with usual care(15). The combined program group had significantly greater weight loss at 12 months (−3.1 kg) than the online program only group (−1.9 kg), and the usual care group (−1.2 kg); these trends persisted at 18 months.

Prior blood pressure telehealth studies have combined remote monitoring with lifestyle modification and antihypertension medication treatment, making it challenging to assess the benefits of lifestyle modification through telehealth/online programs on blood pressure (16, 17). Few data exist on the efficacy of these strategies in rural populations and limited data exists on the benefits of dietitian medical nutrition therapy on blood pressure, assessed by 24-hour ambulatory blood pressure monitoring (ABPM). Use of 24-hour ABPM enhances the power to detect changes in mean blood pressure through repeated measurements while also allowing examination of nocturnal blood pressure, which is associated with increased cardiovascular risk(18). We conducted a randomized controlled trial comparing the effect of two strategies (self-guided vs. a telehealth dietitian-led approach), both using online programs and mobile applications to promote healthy behavior change, on 12-week change in 24-hour systolic blood pressure and other measures of healthy lifestyle. We hypothesized that the telehealth dietitian-led approach would result in a greater decrease in 24-hour systolic BP compared to the self-guided arm.

## Methods

### Overall study design

The Healthy BP study was a parallel arm, randomized clinical trial conducted at 2 hospitals (Geisinger Medical Center, Geisinger Wyoming Valley) in the Geisinger Health System. The study intervention period was 12 weeks to minimize the chance that a participant in the study would undergo a change in antihypertensive medication and because prior studies have shown this timeframe is sufficient to show an improvement in blood pressure through lifestyle changes (2, 19, 20). The ethics committee/Institutional Review Board of Geisinger Medical Center gave ethical approval for this work on 10/2/2018. As described previously in the Study Protocol paper(21), the Novel Coronavirus Disease 2019 (COVID-19) pandemic occurred in the middle of the trial, necessitating protocol adaptations to preserve clinic space, personal protective equipment, and minimize in-person contact. Nevertheless, primary outcome data collection procedures were not altered throughout the trial period.

### Study Participants

Eligibility criteria included: ≥18 years of age, body mass index (BMI) >25 kg/m^2^, access to a telephone and either a computer or smartphone with internet access, 24-hour ambulatory systolic BP between 120-160 mmHg, and successful completion of the run-in period (i.e., enter dietary data for at least 5 out of 7 days and enter weight on the online platform). Exclusion criteria included: inability to understand English, myocardial infarction, stroke, or atherosclerotic cardiovascular disease within prior 6 months, planned or previous bariatric surgery, pregnancy, breast-feeding, or planned pregnancy prior to the end of participation, self-reported average consumption of >21 alcoholic beverages per week or binge drinking, psychiatric hospitalization in past year, current angina, or plans to leave the area prior to the end of the study, current participation in another clinical trial, and principal investigator discretion (i.e. concerns about safety, compliance). Providers were notified by the principal investigator (AC) via electronic message about their patients’ participation in the study. For patients with 24-hour or daytime systolic BP ≥145 mmHg, cases were discussed with patients and their providers on whether to start or escalate antihypertensive medication treatments prior to participation in the study. Participants who had blood pressure medication changes as a result of the 24-hour ABPM were re-evaluated for eligibility at least 2 weeks after a medication change.

### Trial Conduct

Patients with elevated BP were identified primarily through electronic health records, as well as via Geisinger Health Plan health screenings, referrals from primary care providers, and self-referrals. Potential participants were then invited to complete a 24-hour ABPM test to confirm that their BP was elevated outside the office as recommended by guidelines(22). Enrollment began in January 2019 and ended in March 2021. Baseline data were collected during the 24-hour ABPM screening and in a study visit (initially in-person; during the pandemic remote), and follow-up data were collected 12 weeks after randomization **(Figure 1)**.

**Figure 1.**
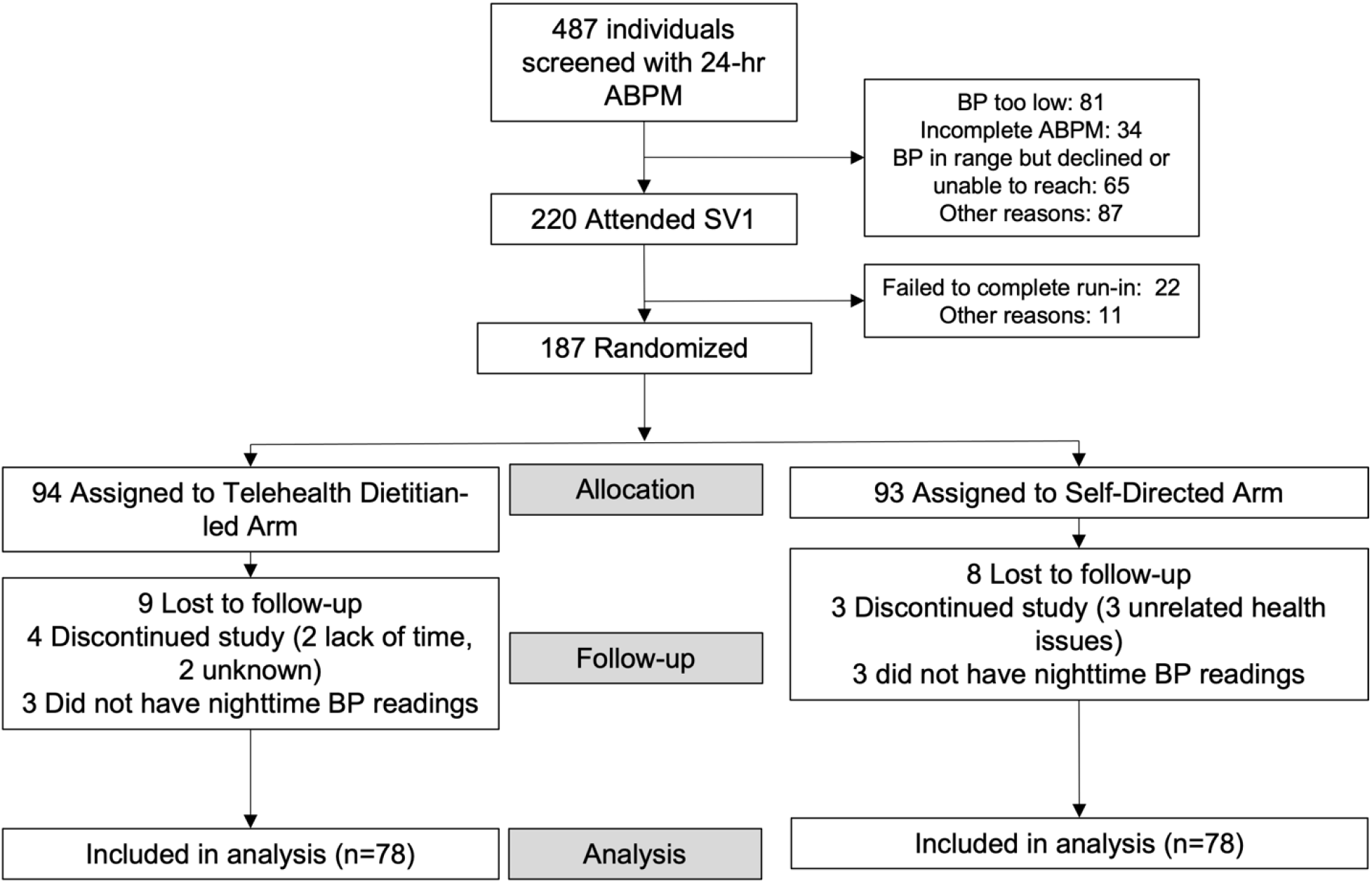
Study Flow Diagram.

### Randomization/Blinding

Randomization assignments were made centrally by a computer program, with an unblinded staff member (not involved in intervention or data collection) opening a sealed opaque envelope with the randomized intervention sequence using a computer program. Participants were notified of their intervention group assignment after completing the run-in period. Eligible participants were randomly assigned 1:1 to two groups: 1) self-guided or 2) dietitian-led telehealth. The outcome assessor remained blinded throughout the study.

### Intervention

All participants received instructions on accessing the Evolve online platform (formerly known as BMIQ)(15, 23). Evolve is an evidence-based, Health Insurance Portability and Accountability Act (HIPAA)-compliant program developed by Dr. Louis Aronne. An earlier version of the platform was used from January 2019 to December 2020 in which participants used a meal-logging app (LoseIt) that integrated with the web platform. The industry-led updates to the platform was phased-in during December 2020 and live through March 2021. The new platform included all of the same elements; meal-logging occurred directly on the Evolve platform rather than requiring the use of an external app. Educational materials on the platform (with adaptations by the study team) included materials to promote weight loss, eating a DASH-type diet (increased intake of fruits/vegetables, whole grains, protein sources from plants, lean meats, and seafood, and reduced intake of sodium, sweets, and saturated fat), increasing physical activity, setting goals, overcoming barriers, and relapse prevention. At baseline, participants received a personalized nutrition report based on a food frequency questionnaire (Viocare) that provided personalized suggestions to improve dietary habits to best align with national dietary guidelines(24). Participants (and their providers) were encouraged to avoid BP medication changes during the 12-week period when possible.

To encourage adherence with dietary data entry, participants received virtual lottery tickets (lottery drawings for ten $300 gift certificates) each week in which they entered at least 5 days/week of dietary data and 2 days/week of weights. All participants were offered the opportunity to participate in grocery store tours at a local grocery store (Weis Markets); after March 2020 grocery store tours were offered virtually.

Overarching participant goals include: 1) weight loss of 3% at 12 weeks; 2) consume a healthier dietary pattern (high in fruits, vegetables, whole grains, low-fat dairy, vegetable/fish/poultry sources of protein, healthier sources of fat, and avoid sweets and salt); 3) reduce sodium intake to <2300 mg/d; 4) at least 180 min/wk of moderate/vigorous intensity physical activity.

### The self-guided intervention group

Participants in this arm received no additional support. Nonclinical study staff monitored participants’ use of the online platform through a web portal and reached out to participants if no data was logged for more than a week via text messages or phone calls to troubleshoot any problems and encourage participation.

### The telehealth dietitian-led intervention group

Participants in the telehealth dietitian arm additionally received weekly telephone calls (initial call 30-60 minutes, follow-up calls 15-20 minutes) from a study dietitian. Our approach was informed by Carver and Scheier’s control systems theory(25) and the situated learning theoretical framework(26). Dietitians received motivational interviewing training bi-annual by an expert (CC). Dietitians examined meal logs of participants with special focus on sodium, fruits, and non-starchy vegetables. Dietitians explored participants’ goals and values and helped participants set attainable goals around a health behavior for the week and assessed their progress towards those goals.

### Data collection

Data collection occurred by telephone and through in-person visits with adaptations during the COVID-19 pandemic (Supplemental Table 1)(22). The enrollment process included a baseline 24-hour SBP between 120-160 mmHg. ABPM was conducted using SpaceLabs Ontrak devices. Participants were asked to record awake and sleep times, and to repeat the 24-hour ABPM if fewer than 14 awake measurements or 7 sleep measurements were obtained. For participants who failed to report awake or sleep times on the activity diary, we visually examined the SpaceLabs Ontrak activity monitor to determine awake and sleep times (27). Prior to March 2020, weight, waist circumference, and automated office blood pressure (AOBP) were measured by research staff using standardized procedures. After March 2020, visits were shifted to mostly remote visits to minimize potential exposure and preserve healthcare resources. Collection of the primary outcome remained unchanged. Study staff collecting 24-hr ABPM data remained blinded to randomization assignment. Weight was self-measured at home by participants, and AOBP was unable to measured during the pandemic. Questionnaires including the web-based Viocare FFQ (24) and the International Physical Activity Questionnaire (IPAQ) Short Form(28) were collected during in-person visits initially; during the pandemic an email with a link to the online FFQ was provided and the IPAQ Short Form was collected by telephone.

### Outcomes

The primary outcome was change from baseline to 12 weeks in 24-hour systolic BP. Secondary outcomes included changes in awake/sleep systolic BP, 24-hour/awake/sleep diastolic BP, total Healthy Eating Index (HEI)-2015 score, weight, and physical activity (total metabolic equivalent of task [MET]-minutes per week). Other outcomes included change in individual components of the HEI-2015 score, end-of-study participant satisfaction, and post-hoc outcomes of sodium/potassium ratio, weight loss >3%, and end-of-study mean awake BP <130/80. Prior to the COVID-19 pandemic, changes in office systolic and diastolic BP were secondary outcomes, assessed by the average of 3 seated office BP readings taken after a 5-minute rest period with 1-minute between measurements; unfortunately, office automated office BP could not be measured after March 2020 due to COVID-19 pandemic restrictions.

### Statistical Analysis

Primary analyses were intention-to-treat including all randomized participants who completed the trial with outcomes data, using unpaired t tests. Complete case analyses were conducted; participants with missing data were excluded from analyses. A 2-sided alpha of 0.05 was used for analyses. We also tested for an interaction between treatment assignment and baseline 24-hour systolic BP (≥ or < 130 mmHg). Since it is possible that the effects of the interventions could be different during the COVID-19 pandemic, we conducted exploratory analyses testing whether change from baseline to 12 weeks in 24-hr systolic BP and other study outcomes differed in participants who completed the intervention prior to 3/1/2020 vs. participants who completed the intervention after 3/1/2020. We conducted a sensitivity analysis, excluding non-adherent participants (< 25% weeks with adequate completed dietary data entry, defined as at least 3 eating occasions ≥ 5 days/week). Outcome measures were assessed for a normal distribution, and non-normally distributed outcomes were transformed as appropriate. Continuous outcomes were compared via linear regression, while dichotomized outcomes were compared via logistic regression.

Based on a sample size of 150 participants completing the study, we estimated that we would have >80% power to detect a difference of 4.6 mmHg between study arms in change of 24-hour systolic BP, a difference of 6.9 units in total HEI-2015 score, and a 2.1 kg difference between groups. All analyses were performed using STATA 15.1 (StataCorp, College Station, TX).

## Results

Out of 487 participants who were assessed for eligibility with 24-hour ABPM, 220 attended a study visit 1, and 187 were randomized **(Figure 1)**. Recruitment was stopped in March 2021 at the discretion of the principal investigator as the study retention rate was higher than expected and because of limited funds. Baseline characteristics were similar between both arms (**Table 1**). Mean age was 54.6 (standard deviation [SD] 13.1) years, mean BMI was 34.5 (6.5) kg/m^2^, 52% were female, 97% were non-Hispanic white, 7% had diabetes, and 25% had depression/anxiety. Baseline mean 24-hr SBP and DBP were 132.7 (8.6) and 77.2 (7.7) mmHg, 67% had daytime BP ≥ 135/85 mmHg, and 26% were on BP medications. A total of 156 (83.4%) completed a follow-up 24-hour ABPM measurement with 78 completers in each arm.

**Table 1.** Baseline Characteristics.

|  | **Self-directed (n=93)** | **Dietitian telehealth (n=94)** |
| --- | --- | --- |
| Age, mean (SD), years | 54.5 (13.0) | 54.8 (13.2) |
| Female | 47 (51%) | 50 (53%) |
| Non-Hispanic white | 90 (97%) | 91 (97%) |
| Education level |  |  |
| Less than high school | 0 (0.0) | 4 (4%) |
| High school | 18 (19%) | 12 (13%) |
| Some college or trade school | 32 (34%) | 40 (43%) |
| College degree | 27 (29%) | 25 (27%) |
| Graduate or Professional school | 16 (17%) | 13 (14%) |
| Annual Household Income |  |  |
| <\$25,000 | 6 (6%) | 6 (6%) |
| \$25-50,000 | 19 (20%) | 22 (23%) |
| \$50-75,000 | 14 (15%) | 18 (19%) |
| \$75,000-100,000 | 15 (16%) | 22 (23%) |
| ≥\$100,000 | 38 (41%) | 25 (27%) |
| Not reported | 0 (0%) | 1 (1%) |
| Employment status |  |  |
| Full-time | 56 (60%) | 52 (55%) |
| Part-time | 12 (13%) | 10 (11%) |
| Unemployed | 3 (3%) | 7 (7%) |
| Retired | 22 (24%) | 23 (24%) |
| Disabled | 0 (0%) | 2 (2%) |
| Smoking status |  |  |
| Current | 4 (4%) | 9 (10%) |
| Former | 19 (20%) | 23 (24%) |
| Never | 69 (74%) | 62 (66%) |
| On BP medications | 24 (25.8%) | 19 (20.2%) |
| Diabetes | 5 (5.4%) | 8 (8.5%) |
| Dyslipidemia | 19 (20.4%) | 29 (30.9%) |
| Depression/anxiety | 24 (26%) | 22 (23%) |
| Body Mass Index, kg/m<sup>2</sup> | 34.8 (6.7) | 34.2 (6.2) |
| Waist circumference, cm | 113.7 (15.3) | 113.2 (14.9) |
| 24-h SBP, mean (SD), mmHg | 133.1 (8.8) | 132.4 (14.9) |
| 24-h DBP, mean (SD), mmHg | 77.1 (8.5) | 77.3 (6.8) |
| Daytime SBP, mean (SD), mmHg | 138.6 (9.0) | 137.9 (8.7) |
| Daytime DBP, mean (SD), mmHg | 81.3 (9.3) | 81.3 (7.2) |
| Nighttime SBP, mean (SD), mmHg | 120.5 (11.3) | 120.2 (10.3) |
| Nighttime DBP, mean (SD), mmHg | 67.5 (8.1) | 68.6 (7.4) |
| HEI-2015 score*, units | 59.7 (11.7) | 61.8 (10.4) |
| MET-minutes per week from moderate/vigorous activity | 1589 (2312) | 1392 (2043) |
| MET-minutes per week from walking | 1137 (1398) | 1006 (1227) |

\*Total HEI-2015 Score range 0-100
Abbreviations: systolic blood pressure (SBP), diastolic blood pressure (DBP) healthy eating index-2015 (HEI-2015), metabolic equivalent of task (MET)

## Intervention Fidelity

Over the 12-week intervention period, the median number of weeks in which dietary data was entered adequately was 10 (interquartile interval 5-12), and the proportion of participants who logged fewer than 25% of weeks was similar in both arms (11.7% in the telehealth dietitian arm, 17.2% in the self-directed arm; p=0.3). Among telehealth dietitian arm participants, the median number of weekly phone calls completed was 11 (IQI 9-12). Only 3 participants (2 telehealth dietitian, 1 self-directed) had an antihypertensive medication change during the study period.

The primary outcome, 24-hr systolic BP decreased from baseline to 12 weeks in both the telehealth dietitian arm (−6.73 mmHg, 95% CI: −8.64, −4.82) and the self-directed arm (−4.92 mmHg, 95% CI: −7.01, −2.77) with no difference between arms (−1.81 mmHg, 95% CI: −4.66, 1.05; p=0.2) (**Table 2, Figure 1)**. However, sleep systolic BP decreased more in the telehealth dietitian arm (−6.92 mmHg, 95% CI: −9.33, −4.51) than the self-directed arm (−1.45 mmHg, 95% CI: −4.28, 1.38) with a difference of −5.47 mmHg (95% CI: −9.16, −1.79; p=0.004) between groups. Similarly, sleep diastolic BP also decreased greater in the telehealth dietitian arm than the self-directed arm (−4.04 mmHg, 95% CI: −6.35, −1.73; p=0.001) (**Table 2, Figure 2)**. Awake systolic and diastolic BP decreased similarly in both arms. At the end of the trial, 41.0% of the telehealth dietitian arm had awake BP <130/80 mmHg and 28.2% in the self-directed arm had awake BP <130/80 mmHg (p=0.09).

**Table 2.** Primary and Secondary Outcomes.

|  | **Telehealth Dietitian Arm** | **Self-directed arm** | **Difference between arms (telehealth – self-directed)** | **P value between groups** |
| --- | --- | --- | --- | --- |
| **24-hr SBP, mmHg (n=156)** | -6.73 (-8.64, -4.82) | -4.92 (-7.01, -2.77) | -1.81 (-4.66, 1.05) | 0.2 |
| **Awake SBP, mmHg (n=156)** | -7.12 (-9.16, -5.07) | -6.21 (-8.28, -4.13) | -0.91 (-3.81, 1.99) | 0.5 |
| **Sleep SBP, mmHg (n=156)** | -6.92 (-9.33, -4.51) | -1.45 (-4.28, 1.38) | -5.47 (-9.16, -1.79) | 0.004 |
| **24-hr DBP, mmHg (n=156)** | -3.46 (-4.67, -2.25) | -2.00 (-3.52, -0.48) | -1.46 (-3.39, 0.47) | 0.1 |
| **Awake DBP, mmHg (n=156)** | -7.62 (-8.79, -6.44) | -6.31 (-7.94, -4.68) | -1.31 (-3.30, 0.69) | 0.2 |
| **Sleep DBP, mmHg (n=156)** | -3.31 (-4.83, -1.78) | 0.73 (-1.03, 2.49) | -4.04 (-6.35, -1.73) | 0.001 |
| **Weight, kg (n=159)** | -5.11 (-6.15, -4.08) | -3.89 (-4.92, -2.86) | -1.22 (-2.68, 0.23) | 0.1 |
| **HEI-2015 score\*, units (n=150)** | 9.23 (6.79, 11.68) | 6.43 (4.30, 8.56) | 2.80 (-0.42, 6.03) | 0.09 |
| **Total MET-mins per week (n=156)** | 866 (250, 1483) | -243 (-869, 382) | 1109 (238, 1981) | 0.01 |
| **AOBP SBP†, mmHg (n=71)** | -6.38 (-10.72, -2.04) | -3.32 (-8.80, 2.16) | -3.05 (-9.86, 3.75) | 0.4 |
| **AOBP DBP†, mmHg (n=71)** | -5.14 (-8.51, -1.76) | -5.24 (-8.99, -1.48) | 0.10 (-4.85, 5.05) | 1.0 |

\*Total HEI-2015 Score range 0-100
†Automated office BP was only taken on participants who completed the study prior to March 2020
Abbreviations: systolic blood pressure (SBP), diastolic blood pressure (DBP) healthy eating index-2015 (HEI-2015), metabolic equivalent of task (MET), healthy eating index-2015 (HEI-2015), automated office blood pressure (AOBP)

**Figure 2.**
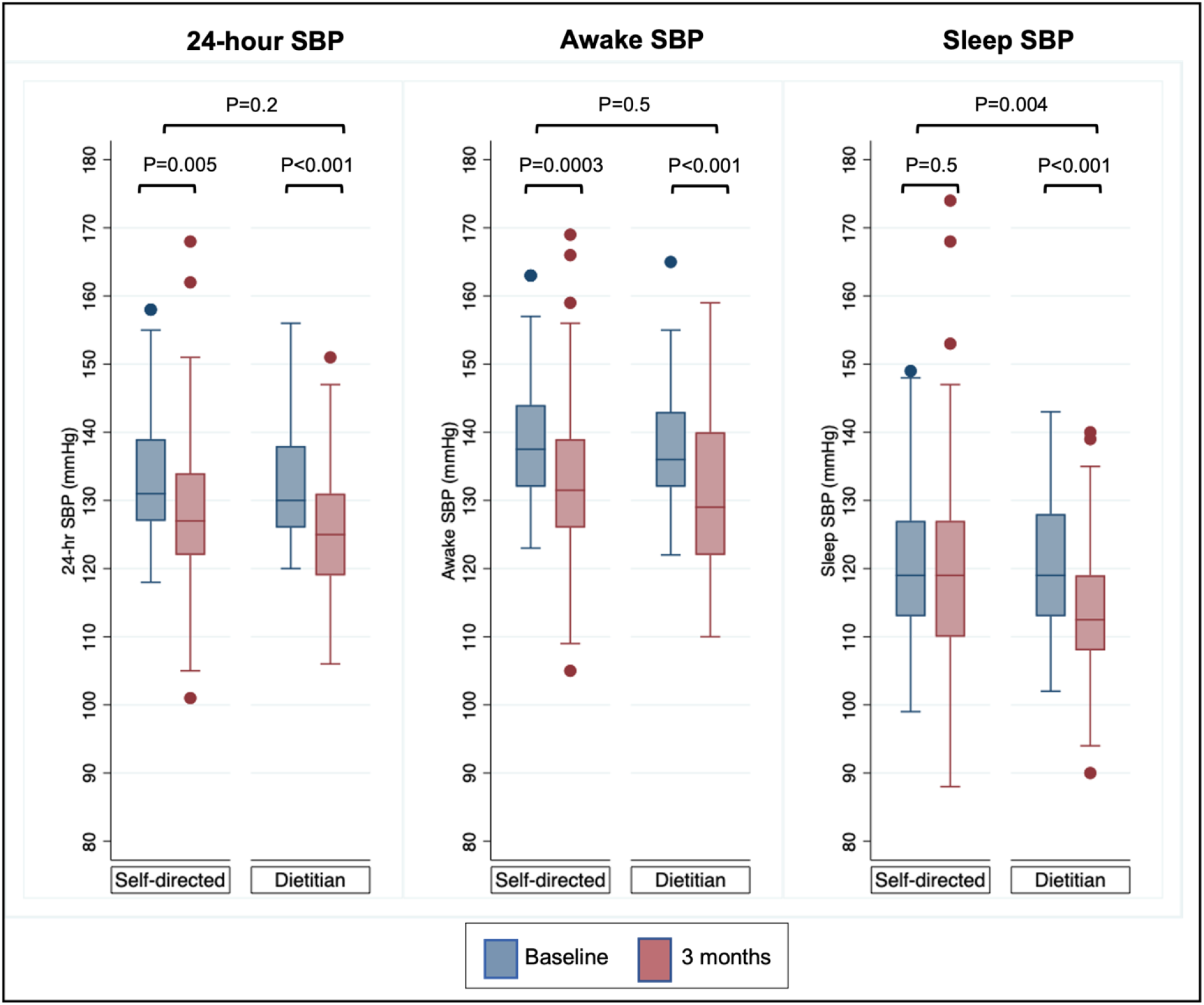
Changes in Systolic Blood Pressure measured by 24-hr ABPM.

The dietitian telehealth arm also had greater improvements in total MET-minutes per week (1109, 95% CI: 238, 1981; p=0.01), with numerically greater but insignificant improvements in HEI-2015 score (2.80, 95% CI: −0.42, 6.03; p=0.09), weight (−1.22 kg, 95% CI: −2.68, 0.23; p= 0.1), and sodium-potassium ratio (− 0.12, 95% CI: −0.28, 0.03; p=0.1), compared to the self-directed arm. Weight loss >3% was achieved by 50.0% in the telehealth dietitian arm and 41.9% in the self-directed arm (p=0.3). The telehealth dietitian arm had greater improvements in HEI-2015 subcategories including total vegetables, greens and beans, total protein foods, and refined grains (**Table 3**). Participants in the telehealth dietitian arm reported greater satisfaction with the research study than the self-directed arm (**Figure 3**).

**Table 3.** Changes in Individual Components of HEI-2015 Score, Sodium, and Potassium, by Randomization Group.

|  | **Dietitian telehealth Arm** | **Self-directed arm** | **Dietitian - Self-directed** | **P value** |
| --- | --- | --- | --- | --- |
| **Total HEI-2015 score** | 9.23 (6.79, 11.68) | 6.43 (4.30, 8.56) | 2.80 (-0.42, 6.03) | 0.09 |
| Total fruits | 1.25 (0.82, 1.68) | 0.99 (0.68, 1.31) | 0.26 (-0.27, 0.79) | 0.3 |
| Whole fruits | 1.15 (0.77, 1.52) | 1.04 (0.72, 1.36) | 0.11 (-0.38, 0.60) | 0.2 |
| Total vegetables | 0.69 (0.48, 0.91) | 0.35 (0.13, 0.57) | 0.34 (0.03, 0.65) | 0.03 |
| Greens and beans | 1.05 (0.74, 1.36) | 0.52 (0.19, 0.84) | 0.54 (0.09, 0.99) | 0.02 |
| Whole grains | 1.06 (0.28, 1.85) | 0.81 (0.13, 1.50) | 0.25 (-0.79, 1.29) | 0.6 |
| Dairy | -0.41 (-0.97, 0.16) | -0.11 (-0.69, 0.47) | -0.30 (-1.09, 0.50) | 0.5 |
| Total protein foods | 0.22 (0.01, 0.43) | -0.08 (-0.30, 0.14) | 0.30 (0.00, 0.60) | 0.05 |
| Seafood and plant proteins | 0.42 (0.12, 0.73) | 0.08 (-0.29, 0.46) | 0.34 (-0.14, 0.82) | 0.2 |
| Fatty acids | 1.45 (0.83, 2.07) | 1.04 (0.48, 1.60) | 0.41 (-0.42, 1.24) | 0.3 |
| Refined grains | 1.09 (0.54, 1.64) | 0.36 (-0.11, 0.83) | 0.73 (0.01, 1.45) | 0.05 |
| Sodium | -0.84 (-1.52, -0.15) | -0.49 (-1.19, 0.22) | -0.35 (-1.33, 0.63) | 0.5 |
| Added sugars | 0.53 (0.13, 0.93) | 0.57 (0.02, 1.12) | -0.04 (-0.71, 0.64) | 0.9 |
| Saturated fats | 1.55 (0.89, 2.21) | 1.34 (0.73, 1.95) | 0.21 (-0.69, 1.10) | 0.6 |
| **Total calories, kcal/d** | -674 (-891, -457) | -546 (-753, -339) | -611 (-759, -462) | 0.4 |
| **Sodium, mg/d** | -920 (-1252, -587) | -845 (-1156, -535) | -74 (-526, 378) | 0.8 |
| **Sodium density, (mg/kcal/d)** | 0.13 (0.04, 0.22) | 0.04 (-0.04, 0.12) | 0.09 (-0.03, 0.21) | 0.1 |
| **Potassium, mg/d** | -307 (-589, -25) | -341 (-585, -97) | 34 (-336, 405) | 0.9 |
| **Potassium density (mg/kcal/d)** | 0.43 (0.33, 0.54) | 0.22 (0.13, 0.31) | 0.21 (0.07, 0.35) | 0.003 |
| **Sodium-potassium molar ratio** | -0.35 (-0.46, -0.23) | -0.22 (-0.33, -0.12) | -0.12 (-0.28, 0.03) | 0.1 |

**Figure 3.**
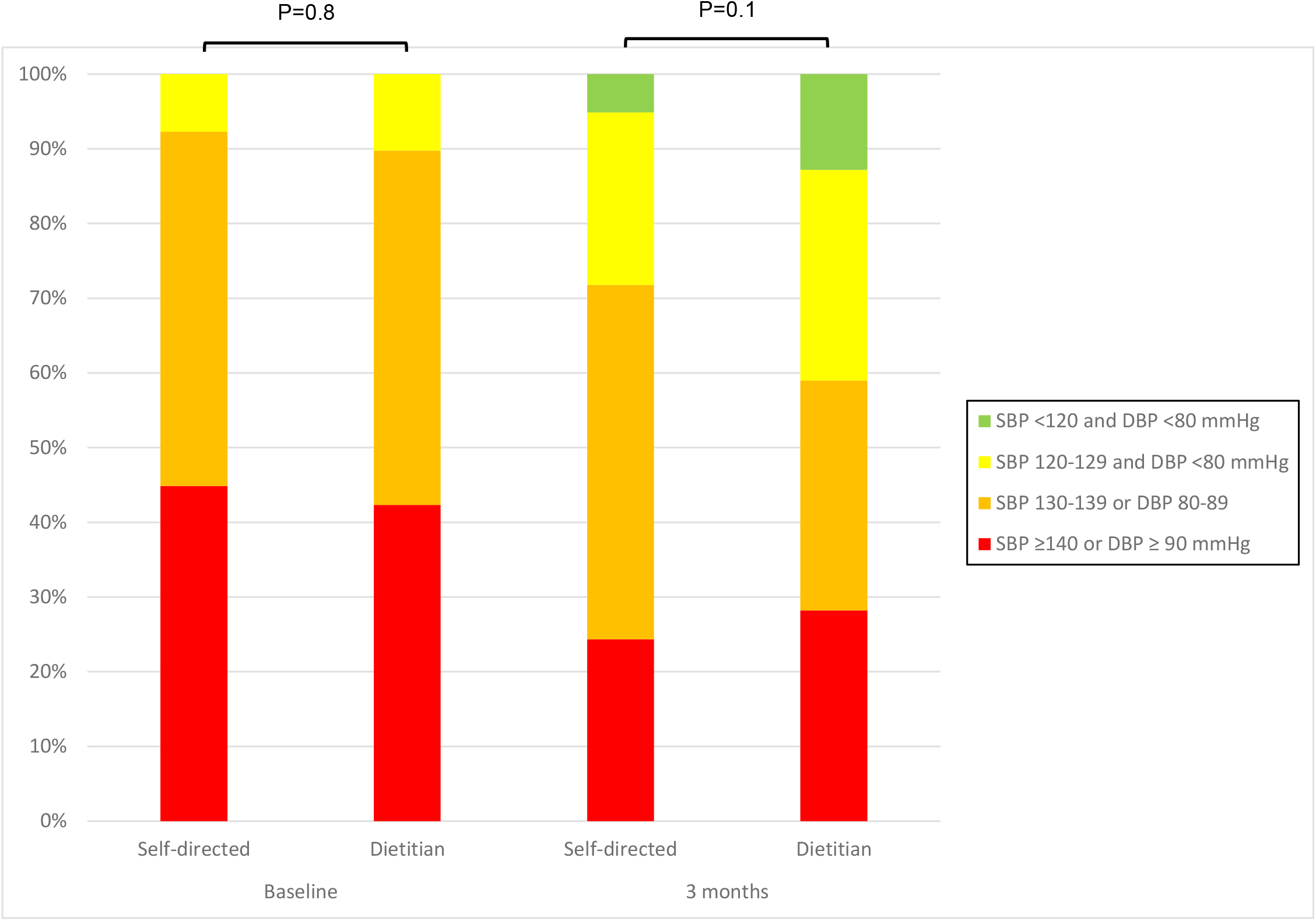
AHA BP Categories at Baseline and 3 months.

**Figure 4.**
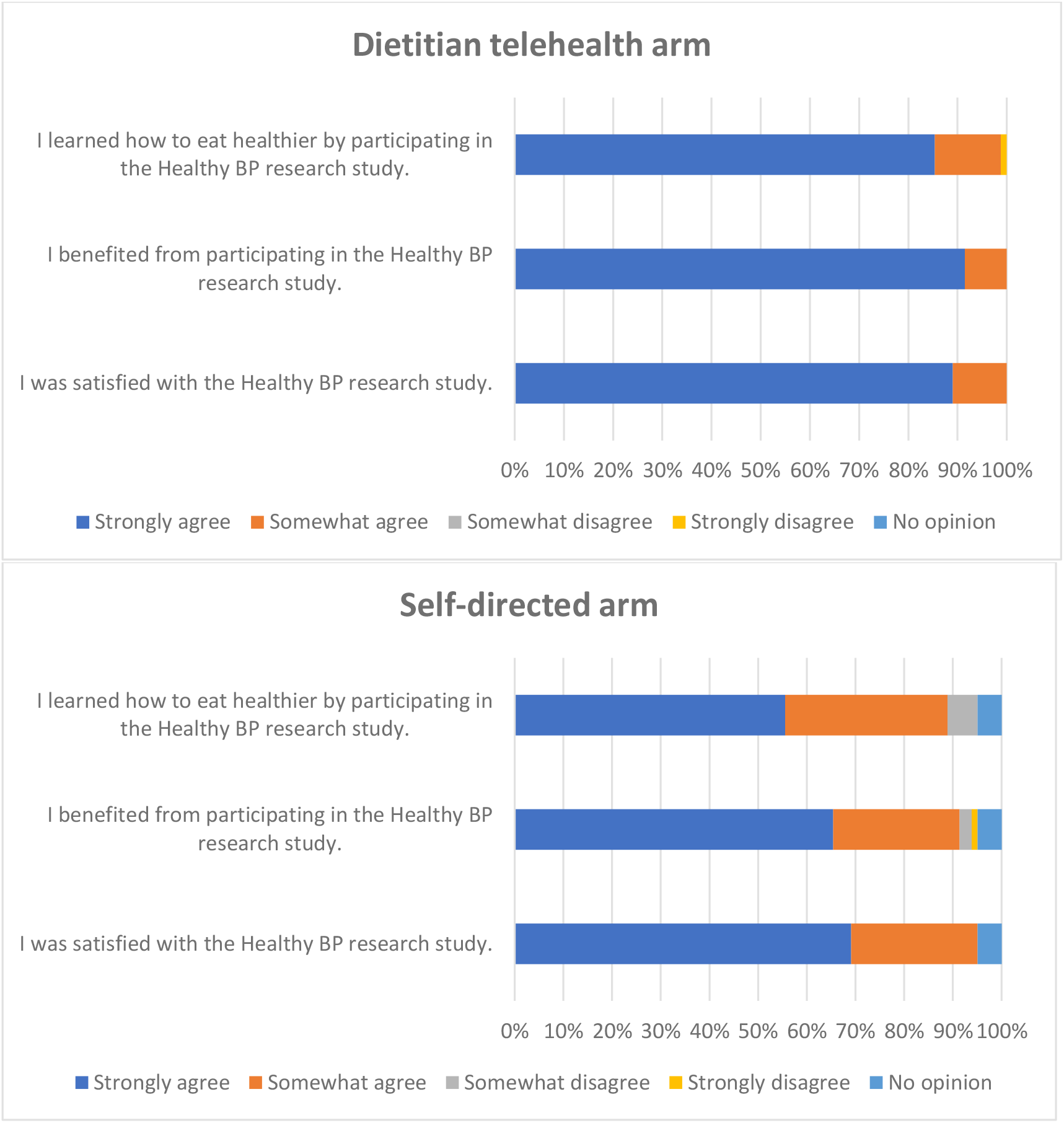
Satisfaction by Study Arm.

There were no significant interactions between treatment assignment and baseline 24-hr systolic BP (≥ or <130 mmHg; p=0.4) or between participation during the COVID-19 pandemic and any of the study outcomes (**Supplemental Table 1**). Among 71 pre-COVID period participants, changes in SBP measured by AOBP were −6.38 (95% CI: −10.72, −2.04) in the telehealth dietitan arm and −3.32 mmHg (95% CI: −8.80, 2.16) in the self-directed arm (**Table 2**). Results were similar in sensitivity analyses excluding 12 participants who logged adequate dietary data <25% of the weeks (data not shown).

There were 3 adverse events unrelated to the study interventions, including 2 new cancer diagnoses and a snow tubing accident resulting in multiple fractures.

## Discussion

In this randomized controlled trial, we demonstrate that both self-directed and telehealth dietitian-led approaches, combined with online programs and web-based apps, resulted in similar reductions in 24-hour systolic BP in adults with overweight/obesity and elevated BP. While there were no significant differences in the primary outcome 24-hr systolic BP, we observed significantly greater reductions in sleep systolic BP and diastolic BP in the telehealth dietitian arm than the self-directed arm. In addition, the telehealth dietitian arm significantly improved physical activity more than the self-directed arm, and tended to improve dietary quality and weight loss.

The observed greater improvements in sleep systolic and diastolic BP in the telehealth dietitian arm are important to note as numerous studies have found that nocturnal BP and a lack of nighttime fall in BP (i.e. “non-dipping”) are important risk factors for cardiovascular disease and mortality, even after adjusting for traditional risk factors and 24-hour systolic BP (18, 29, 30). One explanation for why the telehealth dietitian arm experienced greater improvements in sleep BP but not awake BP could be that measurement during sleep is more standardized (laying down, relatively motionless) than awake BP. Increased physical activity during awake hours of BP measurement could have resulted in transiently higher daytime BP related to physical activity, which was higher in the dietitian telehealth arm than the self-directed arm. In contrast to our findings, Lopes et al. reported that a supervised aerobic training program successfully reduced 24-hr systolic BP and daytime BP with no significant effect on nighttime systolic BP (31). A key difference in their trial was that participants were instructed to not exercise during 24-hr ambulatory BP measurements whereas we instructed participants to continue their regular daily routine without any specific restrictions on exercise. In addition, since 12-week weight loss was excellent in both the telehealth dietitian arm (−5.11 kg) and the self-directed arm (−3.89 kg), the strong effect of weight loss on daytime BP may have overshadowed additional beneficial daytime BP-lowering effects of healthy diet or increased physical activity in the telehealth dietitian group.

The magnitude of the improvements in BP were substantial, similar to a BP medication, and in line with much more resource-intensive feeding studies and lifestyle intervention trials (2, 4, 19, 31). Both the self-directed and the telehealth dietitian arms are scalable interventions that can be easily deployed remotely as the intervention only required internet access and telephone access (no smartphone required). According to the Pew Research Center, 93% of U.S. adults report using the internet and 77% of report having access to home broadband(32). Indeed, the mean age of our participants was 57 with a range from 23 to 89 years of age though our population skewed on the more educated side. While our study did not include a usual care control group, a prior randomized controlled trial found that use of the Evolve platform alone, or in combination with nonclinical staff support, increased weight loss at 12 months (online platform alone −1.9 kg, combined intervention −3.1 kg), compared to usual care (−1.2 kg)(15). Another limitation was that we shifted to remote research visits after March 2022 due to COVID-19 pandemic-related restrictions. This resulted in reliance on participants self-measuring weight and inability to measure AOBP on about half of our study participants. Lastly, our study included a predominantly non-Hispanic white (97%) population, reflecting the demographics of the surrounding, mainly rural area. Additional studies testing the effectiveness of similar remote approaches are needed in more diverse populations.

In conclusion, dietitian-led telehealth and self-directed approaches using web-based applications resulted in similar reductions in 24-hour systolic BP. The dietitian-led intervention lowered nighttime BP and improved physical activity greater than the self-directed arm.

## Data Availability

All data produced in the present study are available upon reasonable request to the authors.

## Acknowledgments

We would like to thank the participants of this research study as well as Dr. Louis Aronne (Evolve) and Rick Weiss (Viocare).

## Data Availability Statement

Data are available upon reasonable request to the authors.

---

**Supplemental Table 1.** Outcomes Pre- and During the COVID-19 Pandemic.

|  |  | Pre-COVID |  |  |  |  | During COVID-19 pandemic |  |  |  |  |
| --- | --- | --- | --- | --- | --- | --- | --- | --- | --- | --- | --- |
|  | N | Telehealth Dietitian Arm | Self-directed arm | Difference between arms (telehealth – self-directed) | P value (between-arm difference) | N | Telehealth Dietitian Arm | Self-directed arm | Difference between arms (telehealth – self-directed) | P value (between-arm difference) | P value for interaction term |
| 24-hr SBP, mmHg | 71 | -5.53 (-8.34, -2.72) | -4.11 (-7.56, -0.67) | -1.41 (-5.77, 2.94) | 0.3 | 85 | -7.77 (-10.44, -5.09) | -5.58 (-8.42, -2.75) | -2.18 (-6.02, 1.66) | 0.3 | 0.8 |
| Awake SBP, mmHg | 71 | -4.92 (-7.80, -2.03) | -5.17 (-8.49, -1.85) | 0.25 (-4.06, 4.57) | 0.9 | 85 | -9.00 (-11.87, -6.13) | -7.05 (-9.78, -4.32) | -1.95 (-5.86, 1.95) | 0.3 | 0.5 |
| Sleep SBP, mmHg | 71 | -7.00 (-10.54, -3.46) | -1.69 (-6.16, 2.79) | -5.31 (-10.90, 0.27) | 0.06 | 85 | -6.86 (-10.29, -3.43) | -1.26 (-5.04, 2.53) | -5.60 (-10.64, -0.56) | 0.03 | 0.9 |
| 24-hr DBP, mmHg | 71 | -2.97 (-4.74, -1.20) | -0.23 (-2.79, 2.33) | -2.74 (-5.79, 0.30) | 0.07 | 85 | -3.88 (-5.59, -2.17) | -3.44 (-5.23, -1.65) | 0.44 (-2.88, 2.01) | 0.7 | 0.2 |
| Awake DBP, mmHg | 71 | -7.39 (-8.93, -5.85) | -4.06 (-6.74, -1.38) | -3.33 (-6.35, -0.32) | 0.03 | 85 | -7.81 (-9.62, -6.69) | -8.14 (-10.05, -6.23) | 0.33 (-2.26, 2.92) | 0.8 | 0.07 |
| Sleep DBP, mmHg | 71 | -3.44 (-5.76, -1.13) | 1.03 (-1.78, 3.84) | -4.47 (-0.90, -8.04) | 0.01 | 85 | -3.19 (-5.30, -1.08) | 0.49 (-1.84, 2.81) | -3.68 (-6.78, -0.58) | 0.02 | 0.7 |
| Weight, kg | 71 | -4.82 (-6.27, -3.37) | -4.74 (-6.44, -3.04) | -0.08 (-2.27, 2.11) | 0.9 | 88 | -5.36 (-6.88, -3.83) | -3.23 (-4.53, -1.93) | -2.13 (-4.10, -0.16) | 0.03 | 0.2 |
| HEI-2015 score, units | 71 | 10.10 (6.10, 14.10) | 6.63 (3.35, 9.91) | 3.47 (-1.62, 8.57) | 0.2 | 83 | 8.49 (5.37, 11.60) | 6.26 (3.33, 9.19) | 2.22 (-1.99, 6.44) | 0.3 | 0.8 |
| Total MET-mins per week | 71 | 598 (217, 1414) | -344 (-1257, 569) | 943 (-258, 2143) | 0.1 | 91 | 1080 (158, 2002) | -167 (-1051, 718) | 1247 (-1089, 190) | 0.05 | 0.8 |
| AOBP SBP†, mmHg | 71 | -6.38 (-10.72, -2.04) | -3.32 (-8.80, 2.16) | -3.05 (-9.86, 3.75) | 0.4 | 0 | N/A | N/A | N/A | N/A | N/A |
| AOBP DBP†, mmHg | 71 | -5.14 (-8.51, -1.76) | -5.24 (-8.99, -1.48) | 0.10 (-4.85, 5.05) | 1.0 | 0 | N/A | N/A | N/A | N/A | N/A |

